# Adverse childhood experiences and resilience among adult women: a population-based study

**DOI:** 10.1101/2021.07.05.21260008

**Authors:** Hilda Björk Daníelsdóttir, Thor Aspelund, Edda Björk Þórðardóttir, Katja Fall, Fang Fang, Gunnar Tómasson, Harpa Rúnarsdóttir, Qian Yang, Karmel W. Choi, Beatrice Kennedy, Thorhildur Halldorsdottir, Donghao Lu, Huan Song, Jóhanna Jakobsdóttir, Arna Hauksdóttir, Unnur Anna Valdimarsdóttir

## Abstract

Adverse childhood experiences (ACEs) have consistently been associated with elevated risk of multiple adverse health outcomes, yet their contribution to coping ability and psychiatric resilience in adulthood is unclear. Participants were 19,613 women in the Icelandic Stress-And-Gene-Analysis cohort with complete data on 13 ACEs measured with the ACE-International Questionnaire. Self-reported coping ability was measured with the Connor-Davidson Resilience Scale and psychiatric resilience was operationalized as absence of psychiatric morbidity. Compared to women with 0 ACEs, women with ≥ 5 ACEs had 33% lower prevalence of high coping ability (PR=0.67, 95% CI 0.60,0.74) and 56% lower prevalence of high psychiatric resilience (PR=0.44; 95% CI 0.41,0.48). Specific ACEs including emotional neglect, bullying, sexual abuse and mental illness of household member were consistently associated with reduced adult resilience. We observed only slightly attenuated associations after controlling for adult socioeconomic factors and social support in adulthood, indicating that adult resilience may be largely determined in childhood.

## Introduction

Exposure to adverse childhood experiences (ACEs), including child abuse, neglect and growing up in dysfunctional households, is associated with elevated risk of a wide range of physical and mental health problems across the life course (Anda et al., 2006; Bellis et al., 2015; Hughes et al., 2017; Petruccelli, Davis, & Berman, 2019). The results from a recent meta-analysis (Hughes et al., 2017) suggest that the adult health outcomes most clearly associated with ACEs include problematic alcohol consumption and substance use, violence, and mental illness. However, there is great variation in long-term outcomes of children exposed to ACEs, and many children remain healthy despite excessive ACE exposure. Importantly, it has been documented that a substantial proportion of individuals exposed to ACEs do not develop mental illness in adulthood (Green et al., 2010; Kessler et al., 2010), but instead exhibit resilience (DuMont, Widom, & Czaja, 2007; Holmes, Yoon, Voith, Kobulsky, & Steigerwald, 2015).

Resilience is generally conceptualized as maintained mental health or positive adaptation despite trauma exposure (Choi, Stein, Dunn, Koenen, & Smoller, 2019; Kalisch et al., 2017; Luthar, Cicchetti, & Becker, 2000). The two most widely used approaches to operationalize the concept define resilience as *perceived coping ability* reflecting a stable tendency to cope effectively with stress and adversity (Campbell-Sills & Stein, 2007; Connor & Davidson, 2003), and as *psychiatric resilience* reflecting an empirically derived outcome, such as the absence of PTSD or other psychiatric disorders among individuals exposed to adversity (Nishimi, Choi, Cerutti, et al., 2020; Sheerin, Stratton, Amstadter, The VA Mid-Atlantic Mental Illness Research, Education, Clinical Center (MIRECC) Workgroup., & McDonald, 2018). The different resilience definitions are not mutually exclusive, but rather complementary and may capture different underlying dimensions of resilience (Choi et al., 2019; Fisher & Law, 2021; Sheerin et al., 2018).

Only a handful of previous studies have addressed the association between ACEs and adult resilience. Childhood maltreatment, variously defined, has been negatively associated with psychiatric resilience (McGloin & Widom, 2001; Mersky & Topitzes, 2010; Topitzes, Mersky, Dezen, & Reynolds, 2013; Williams, MacMillan, & Jamieson, 2006) and perceived coping ability (Campbell-Sills, Forde, & Stein, 2009; Nishimi, Choi, Davis, et al., 2020) in adulthood. However, most studies have focused solely on childhood maltreatment or included only a small number of ACEs. Therefore, to date, little is known about the association between cumulative ACE exposure and adult resilience, and whether specific ACE types are to a varying extent associated with resilience.

Leveraging a large nationwide study of Icelandic women, we aimed to investigate the association between the cumulative number of a broad spectrum of ACEs and two distinct measures of adult resilience, i.e. perceived coping ability and an outcome-based measure of low psychiatric morbidity.

## Material and Methods

### Study sample

In this study, we utilized data from the Stress-And-Gene-Analysis (SAGA) cohort, a population-based study in Iceland on the impact of trauma on women’s health. All 18 to 69-year-old Icelandic speaking women residing in Iceland with an identifiable address or telephone number (n≈104,197), were invited to participate in the study from March 2018. Data collection continued until July 1^st^ 2019, yielding a total of 30,372 participating women (30% of eligible women). Since trauma exposure is intrinsic to psychiatric resilience, the analytic sample was restricted to women reporting a worst traumatic event at some point during their lifetime (see description of PCL-5 below). In addition, women who had missing or incomplete data on ACEs (n=2939), perceived coping ability (n=511), and/or psychiatric resilience (n=4737), were excluded, which resulted in a final study population of 19,613 women (Supplementary Figure 1).

### Measures

#### Adverse childhood experiences (ACEs)

ACEs were measured with a modified version of the Adverse Childhood Experiences International Questionnaire (ACE-IQ) developed by the WHO (‘WHO | ACE-IQ’). The instrument consists of 30 items assessing how often individuals were exposed to the following 13 ACEs during the first 18 years of their life: emotional neglect, physical neglect, emotional abuse, physical abuse, sexual abuse, domestic violence, living with a household member who abuses drugs and/or alcohol, living with a household member who is mentally ill or suicidal, incarceration of a household member, parental death or separation/divorce, being bullied, witnessing community violence, and exposure to war/collective violence. For an overview of included items and their response options see Supplementary Table 1. The recommended frequency scoring system (‘WHO | ACE-IQ’), which takes into account the level of exposure for each ACE, was used to generate three types of exposure variables: 1) a continuous ACE-IQ total score ranging from 0 to 13, reflecting the number of ACEs participants were exposed to; 2) the total score was categorized (0, 1, 2, 3-4 and ≥ 5 ACEs) based on the distribution of the sample; 3) binary variables for each individual ACE type (described above), coded as 0 (unexposed) and 1 (exposed).

#### Perceived coping ability

Perceived coping ability was assessed with the 10-item version of the Connor-Davidson Resilience Scale (CD-RISC-10) (Campbell-Sills & Stein, 2007). The scale has demonstrated good reliability and validity (Campbell-Sills & Stein, 2007). All items were answered on a 5-point scale ranging from 0 (not true at all) to 4 (true nearly all the time). Items were summed to create a total score ranging from 0 to 40 with a higher score indicating higher levels of perceived coping ability. As there is no standardized cut-off for the CD-RISC, the total scores were divided into quintiles (for more details see Supplementary Materials), and a binary variable was created where the highest quintile was used to define a high level of perceived coping ability (i.e. resilience, CD-RISC score ≥ 35).

#### Psychiatric resilience

Consistent with previous literature (Choi et al., 2019; Nishimi, Choi, Davis, et al., 2020; Stein et al., 2019) psychiatric resilience was defined as absence of or low psychiatric morbidity among women exposed to lifetime trauma, i.e. total sum of the inverse number of above-threshold symptom levels on PTSD, trauma-related sleep disturbances, binge drinking, depression, and anxiety.

The PTSD Checklist for DSM-5 (PCL-5) is a 20-item valid instrument that assesses symptoms of PTSD in the past month according to the DSM-5 (Blevins, Weathers, Davis, Witte, & Domino, 2015; Bovin et al., 2016; Weathers et al., 2013; Wortmann et al., 2016). Before answering the PCL-5, participants reported their lifetime exposure to potentially traumatic events assessed with the Life Events Checklist for DSM-5 (LEC-5). Participants were asked to determine the worst traumatic event they had experienced and answer the PCL-5 according to the worst selected trauma. A clinical cut-off score of ≥33 was used to indicate probable PTSD (Weathers et al., 2013). The Pittsburgh Sleep Quality Index Addendum for PTSD (PSQI-A) is a 7-item valid questionnaire designed to assess the frequency of disruptive nocturnal behaviours common in individuals with PTSD (Germain, Hall, Krakow, Katherine Shear, & Buysse, 2005). A clinical cut-off score of ≥4 was used to discriminate between participants with and without trauma-related sleep disturbances (Germain et al., 2005). Binge drinking was defined as having 6 or more units of alcohol on a single occasion (one unit corresponds to a single measure of spirits) at least once a month during the last year (Bush, Kivlahan, McDonell, Fihn, & Bradley, 1998). The widely used Patient Health Questionnaire (PHQ-9) (Kroenke, Spitzer, & Williams, 2001) and Generalized Anxiety Disorder Scale (GAD-7) (Kroenke, Spitzer, Williams, & Löwe, 2010; Löwe et al., 2008; Spitzer, Kroenke, Williams, & Löwe, 2006) were used to measure the presence of depression and anxiety symptoms, respectively, during the past two weeks. The standard cut-off score of ≥10 was used, indicating clinically relevant depression and anxiety (Kroenke et al., 2001; Spitzer et al., 2006).

Binary variables were created indicating whether an individual met the clinical cut-off for symptoms of each disorder above (0=no, 1=yes). The psychiatric resilience phenotype was derived by summing together the binary variables and reversing the score which resulted in a total score ranging from 0 to 5 where higher scores indicate greater psychiatric resilience in these trauma-exposed women. In addition, we created a binary variable where endorsement of 0 on all measures indicated high psychiatric resilience.

#### Covariates

Variables with a conceptual rationale for being associated both with ACEs and perceived coping ability or psychiatric resilience were considered as covariates. These included age (at responding), childhood deprivation, as well as educational level, employment status, civil status and current monthly income at responding. In addition, current perceived social support, measured with the Multidimensional Scale of Perceived Social Support (MSPSS) (Zimet, Dahlem, Zimet, & Farley, 1988), and perceived happiness, measured with a 10-point visual digital scale, were included in additional analyses. For more detailed description of covariate assessment, see the Supplementary Materials.

### Statistical analysis

Descriptive characteristics were compared using Chi-square tests for categorical data and ANOVAs for continuous data.

Rank order correlations were used to determine i) the correlation between perceived coping ability and psychiatric resilience, ii) correlations between perceived coping ability and different measures of psychopathology (PHQ-9, GAD-7, PCL-5, PSQI-A, binge drinking) used to derive psychiatric resilience, and iii) correlations between different ACE subtypes.

We used linear regression models assuming normally distributed errors and log-linear Poisson regression models with robust error variance to determine the associations between ACEs and perceived coping ability and psychiatric resilience, as continuous and binary outcomes (high perceived coping ability/ high psychiatric resilience), respectively. In all analyses, we adjusted for age and childhood deprivation (model 1) and then additionally for adult educational level, civil status, employment level and income (model 2).

We ran all models with ACE-IQ as a continuous predictor and as a categorical predictor where we compared resilience levels of unexposed women (0 ACEs) to resilience levels of those who had been exposed to increasing number of ACEs (1, 2, 3-4 and ≥ 5 ACEs). In addition, we carried out stratified analyses to assess whether the association between ACEs and resilience differed by levels of perceived social support (linear models). Furthermore, because parental separation/divorce is a common childhood experience, we carried out a sensitivity analysis excluding this item from the ACE-IQ score and re-ran the linear models. Finally, to preclude whether a particular response style influenced the observed associations between ACEs and resilience, we excluded women who scored low or high on happiness (10% top/bottom scores) and re-ran the linear models.

To determine the independent associations of specific types of ACEs with resilience, we ran analyses on perceived coping ability and psychiatric resilience for each of the 13 ACEs. We first examined each ACE type separately while adjusting for covariates, and then re-ran the analyses with all ACE subtypes entered simultaneously into the model, as ACEs frequently co-occur (Radford, Corral, Bradley, & Fisher, 2013).

Standardized regression coefficients were reported from all linear regression analyses and prevalence ratios (PR) were reported from the Poisson regression analyses. All statistical analyses were performed using R (version 3.6.1).

## Results

### Characteristics of the sample

Descriptive statistics of the study population are presented in Table 1. Overall, about 22% of participants reported no ACEs and 17% reported five or more ACEs. Middle aged women had on average higher scores on the ACE-IQ than younger or older women. Women with higher ACE scores were more often single or widowed, less educated, unemployed, had lower income and were more likely to report childhood deprivation and low perceived social support (Table 1. *Bivariate associations*

**Table 1.**
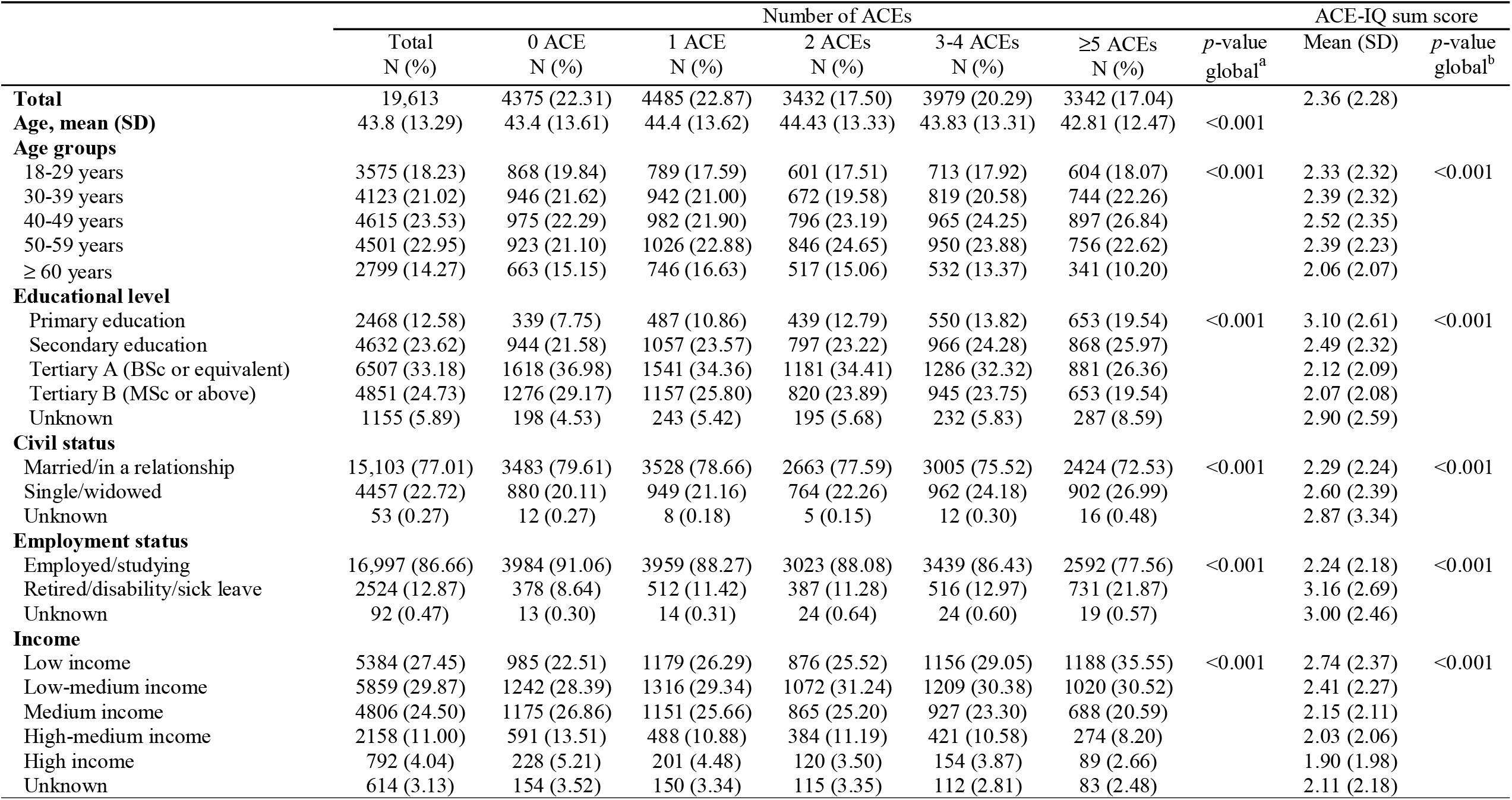
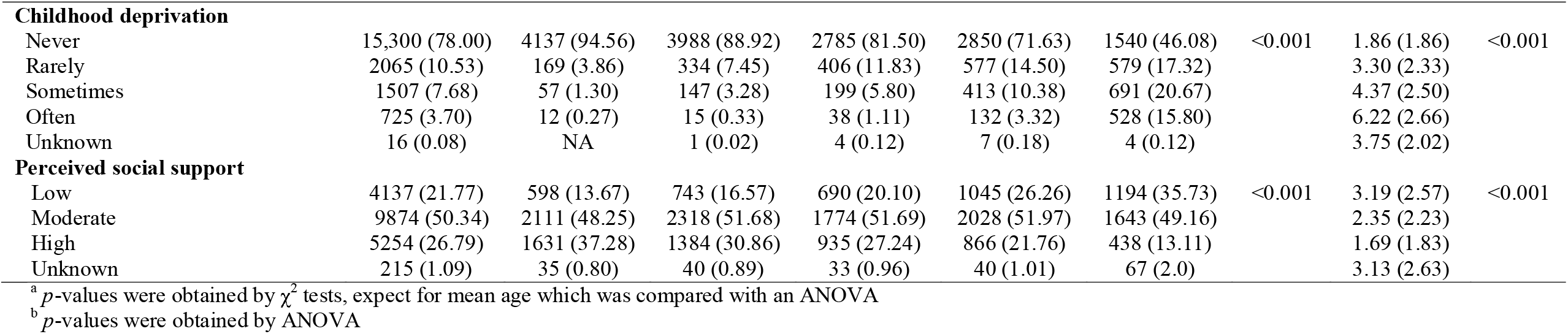
Descriptive characteristics of the study population by number of adverse childhood experiences (ACE-IQ) (n = 19,613)

|  | Number of ACEs |  |  |  |  |  | *p*-value<br>global<sup>a</sup> | ACE-IQ sum score |  |
| --- | --- | --- | --- | --- | --- | --- | --- | --- | --- |
|  | Total<br>N (%) | 0 ACE<br>N (%) | 1 ACE<br>N (%) | 2 ACEs<br>N (%) | 3-4 ACEs<br>N (%) | ≥5 ACEs<br>N (%) |  | Mean (SD) | *p*-value<br>global<sup>b</sup> |
| **Total** | 19,613 | 4375 (22.31) | 4485 (22.87) | 3432 (17.50) | 3979 (20.29) | 3342 (17.04) |  | 2.36 (2.28) |  |
| **Age, mean (SD)** | 43.8 (13.29) | 43.4 (13.61) | 44.4 (13.62) | 44.43 (13.33) | 43.83 (13.31) | 42.81 (12.47) | <0.001 |  |  |
| **Age groups** |  |  |  |  |  |  |  |  |  |
| 18-29 years | 3575 (18.23) | 868 (19.84) | 789 (17.59) | 601 (17.51) | 713 (17.92) | 604 (18.07) | <0.001 | 2.33 (2.32) | <0.001 |
| 30-39 years | 4123 (21.02) | 946 (21.62) | 942 (21.00) | 672 (19.58) | 819 (20.58) | 744 (22.26) |  | 2.39 (2.32) |  |
| 40-49 years | 4615 (23.53) | 975 (22.29) | 982 (21.90) | 796 (23.19) | 965 (24.25) | 897 (26.84) |  | 2.52 (2.35) |  |
| 50-59 years | 4501 (22.95) | 923 (21.10) | 1026 (22.88) | 846 (24.65) | 950 (23.88) | 756 (22.62) |  | 2.39 (2.23) |  |
| ≥ 60 years | 2799 (14.27) | 663 (15.15) | 746 (16.63) | 517 (15.06) | 532 (13.37) | 341 (10.20) |  | 2.06 (2.07) |  |
| **Educational level** |  |  |  |  |  |  |  |  |  |
| Primary education | 2468 (12.58) | 339 (7.75) | 487 (10.86) | 439 (12.79) | 550 (13.82) | 653 (19.54) | <0.001 | 3.10 (2.61) | <0.001 |
| Secondary education | 4632 (23.62) | 944 (21.58) | 1057 (23.57) | 797 (23.22) | 966 (24.28) | 868 (25.97) |  | 2.49 (2.32) |  |
| Tertiary A (BSc or equivalent) | 6507 (33.18) | 1618 (36.98) | 1541 (34.36) | 1181 (34.41) | 1286 (32.32) | 881 (26.36) |  | 2.12 (2.09) |  |
| Tertiary B (MSc or above) | 4851 (24.73) | 1276 (29.17) | 1157 (25.80) | 820 (23.89) | 945 (23.75) | 653 (19.54) |  | 2.07 (2.08) |  |
| Unknown | 1155 (5.89) | 198 (4.53) | 243 (5.42) | 195 (5.68) | 232 (5.83) | 287 (8.59) |  | 2.90 (2.59) |  |
| **Civil status** |  |  |  |  |  |  |  |  |  |
| Married/in a relationship | 15,103 (77.01) | 3483 (79.61) | 3528 (78.66) | 2663 (77.59) | 3005 (75.52) | 2424 (72.53) | <0.001 | 2.29 (2.24) | <0.001 |
| Single/widowed | 4457 (22.72) | 880 (20.11) | 949 (21.16) | 764 (22.26) | 962 (24.18) | 902 (26.99) |  | 2.60 (2.39) |  |
| Unknown | 53 (0.27) | 12 (0.27) | 8 (0.18) | 5 (0.15) | 12 (0.30) | 16 (0.48) |  | 2.87 (3.34) |  |
| **Employment status** |  |  |  |  |  |  |  |  |  |
| Employed/studying | 16,997 (86.66) | 3984 (91.06) | 3959 (88.27) | 3023 (88.08) | 3439 (86.43) | 2592 (77.56) | <0.001 | 2.24 (2.18) | <0.001 |
| Retired/disability/sick leave | 2524 (12.87) | 378 (8.64) | 512 (11.42) | 387 (11.28) | 516 (12.97) | 731 (21.87) |  | 3.16 (2.69) |  |
| Unknown | 92 (0.47) | 13 (0.30) | 14 (0.31) | 24 (0.64) | 24 (0.60) | 19 (0.57) |  | 3.00 (2.46) |  |
| **Income** |  |  |  |  |  |  |  |  |  |
| Low income | 5384 (27.45) | 985 (22.51) | 1179 (26.29) | 876 (25.52) | 1156 (29.05) | 1188 (35.55) | <0.001 | 2.74 (2.37) | <0.001 |
| Low-medium income | 5859 (29.87) | 1242 (28.39) | 1316 (29.34) | 1072 (31.24) | 1209 (30.38) | 1020 (30.52) |  | 2.41 (2.27) |  |
| Medium income | 4806 (24.50) | 1175 (26.86) | 1151 (25.66) | 865 (25.20) | 927 (23.30) | 688 (20.59) |  | 2.15 (2.11) |  |
| High-medium income | 2158 (11.00) | 591 (13.51) | 488 (10.88) | 384 (11.19) | 421 (10.58) | 274 (8.20) |  | 2.03 (2.06) |  |
| High income | 792 (4.04) | 228 (5.21) | 201 (4.48) | 120 (3.50) | 154 (3.87) | 89 (2.66) |  | 1.90 (1.98) |  |
| Unknown | 614 (3.13) | 154 (3.52) | 150 (3.34) | 115 (3.35) | 112 (2.81) | 83 (2.48) |  | 2.11 (2.18) |  |

Table 1 (continued)
|  |  |  |  |  |  |  |  |  |  |
| --- | --- | --- | --- | --- | --- | --- | --- | --- | --- |
| **Childhood deprivation** |  |  |  |  |  |  |  |  |  |
| Never | 15,300 (78.00) | 4137 (94.56) | 3988 (88.92) | 2785 (81.50) | 2850 (71.63) | 1540 (46.08) | <0.001 | 1.86 (1.86) | <0.001 |
| Rarely | 2065 (10.53) | 169 (3.86) | 334 (7.45) | 406 (11.83) | 577 (14.50) | 579 (17.32) |  | 3.30 (2.33) |  |
| Sometimes | 1507 (7.68) | 57 (1.30) | 147 (3.28) | 199 (5.80) | 413 (10.38) | 691 (20.67) |  | 4.37 (2.50) |  |
| Often | 725 (3.70) | 12 (0.27) | 15 (0.33) | 38 (1.11) | 132 (3.32) | 528 (15.80) |  | 6.22 (2.66) |  |
| Unknown | 16 (0.08) | NA | 1 (0.02) | 4 (0.12) | 7 (0.18) | 4 (0.12) |  | 3.75 (2.02) |  |
| **Perceived social support** |  |  |  |  |  |  |  |  |  |
| Low | 4137 (21.77) | 598 (13.67) | 743 (16.57) | 690 (20.10) | 1045 (26.26) | 1194 (35.73) | <0.001 | 3.19 (2.57) | <0.001 |
| Moderate | 9874 (50.34) | 2111 (48.25) | 2318 (51.68) | 1774 (51.69) | 2028 (51.97) | 1643 (49.16) |  | 2.35 (2.23) |  |
| High | 5254 (26.79) | 1631 (37.28) | 1384 (30.86) | 935 (27.24) | 866 (21.76) | 438 (13.11) |  | 1.69 (1.83) |  |
| Unknown | 215 (1.09) | 35 (0.80) | 40 (0.89) | 33 (0.96) | 40 (1.01) | 67 (2.0) |  | 3.13 (2.63) |  |

<sup>a</sup> $p$ -values were obtained by $\chi^2$ tests, except for mean age which was compared with an ANOVA
<sup>b</sup> $p$ -values were obtained by ANOVA

Both perceived coping ability and psychiatric resilience were consistently associated with increased age, higher educational level, being in a relationship or married, being employed, higher income, lower childhood deprivation and higher perceived social support (Table 2). The two resilience measures were moderately correlated (r_s_=0.45). Perceived coping ability was moderately negatively correlated with each of the measures used to derive the psychiatric resilience phenotype (i.e. symptoms of depression, anxiety, PTSD, and trauma-related sleep disturbances) but weakly correlated with binge drinking (Supplementary Table 2). The 13 ACE subtypes were weakly to moderately correlated with each other (r_s_ ranging from 0.05 to 0.46), with emotional abuse showing the strongest correlation with other ACEs (Supplementary Table 3).

**Table 2.** Distribution of perceived coping ability (CD-RISC) and psychiatric resilience scores by sociodemographic characteristics

|  | Perceived coping ability |  | Psychiatric resilience |  |
| --- | --- | --- | --- | --- |
|  | Mean (SD) | *p*-value global<sup>a</sup> | Mean (SD) | *p*-value global<sup>a</sup> |
| **Total** | 28.05 (7.35) |  | 3.76 (1.44) |  |
| **Age groups** |  |  |  |  |
| 18-29 years | 25.54 (7.71) | <0.001 | 3.27 (1.59) | <0.001 |
| 30-39 years | 27.55 (7.38) |  | 3.67 (1.50) |  |
| 40-49 years | 28.37 (7.25) |  | 3.79 (1.42) |  |
| 50-59 years | 29.24 (7.01) |  | 3.95 (1.29) |  |
| ≥ 60 years | 29.53 (6.64) |  | 4.20 (1.17) |  |
| **Educational level** |  |  |  |  |
| Primary education | 24.86 (8.22) | <0.001 | 3.23 (1.57) | <0.001 |
| Secondary education | 26.78 (7.55) |  | 3.58 (1.44) |  |
| Tertiary A (BSc or equivalent) | 28.55 (6.90) |  | 3.88 (1.33) |  |
| Tertiary B (MSc or above) | 30.28 (6.37) |  | 4.10 (1.21) |  |
| Unknown | 27.72 (7.50) |  | 3.63 (1.43) |  |
| **Civil status** |  |  |  |  |
| Married/in a relationship | 28.31 (7.21) | <0.001 | 3.86 (1.39) | <0.001 |
| Single/widowed | 27.17 (7.75) |  | 3.44 (1.55) |  |
| Unknown | 25.47 (8.29) |  | 3.08 (1.81) |  |
| **Employment status** |  |  |  |  |
| Employed/studying | 28.57 (7.03) | <0.001 | 3.86 (1.38) | <0.001 |
| Retired/disability/sick leave | 24.68 (8.42) |  | 3.14 (1.60) |  |
| Unknown | 23.96 (9.19) |  | 3.20 (1.69) |  |
| **Income** |  |  |  |  |
| Low income | 25.23 (7.83) | <0.001 | 3.28 (1.58) | <0.001 |
| Low-medium income | 27.58 (7.13) |  | 3.76 (1.43) |  |
| Medium income | 29.65 (6.43) |  | 4.06 (1.27) |  |
| High-medium income | 31.30 (5.92) |  | 4.14 (1.21) |  |
| High income | 32.39 (5.78) |  | 4.22 (1.11) |  |
| Unknown | 27.65 (7.83) |  | 3.91 (1.37) |  |
| **Childhood deprivation** |  |  |  |  |
| Never | 28.48 (7.17) | <0.001 | 3.92 (1.36) | <0.001 |
| Rarely | 27.19 (7.43) |  | 3.44 (1.53) |  |
| Sometimes | 26.18 (7.83) |  | 3.19 (1.57) |  |
| Often | 25.37 (8.48) |  | 2.72 (1.61) |  |
| Unknown | 22.38 (8.82) |  | 2.19 (1.68) |  |
| **Perceived social support** |  |  |  |  |
| Low | 25.19 (7.96) | <0.001 | 3.21 (1.59) | <0.001 |
| Moderate | 27.64 (7.00) |  | 3.77 (1.42) |  |
| High | 31.22 (6.24) |  | 4.21 (1.15) |  |
| Unknown | 25.73 (7.44) |  | 3.29 (1.57) |  |

<sup>a</sup> *p*-values were obtained by ANOVAs

### Bivariate associations

Both perceived coping ability and psychiatric resilience were consistently associated with increased age, higher educational level, being in a relationship or married, being employed, higher income, lower childhood deprivation and higher perceived social support (Table 2). The two resilience measures were moderately correlated (r_s_=0.45). Perceived coping ability was moderately negatively correlated with each of the measures used to derive the psychiatric resilience phenotype (i.e. symptoms of depression, anxiety, PTSD, and trauma-related sleep disturbances) but weakly correlated with binge drinking (Supplementary Table 2). The 13 ACE subtypes were weakly to moderately correlated with each other (r_s_ ranging from 0.05 to 0.46), with emotional abuse showing the strongest correlation with other ACEs (Supplementary Table 3).

### Associations between ACEs and resilience

Linear models revealed that exposure to one or more ACEs (relative to 0 ACEs) was associated with attenuated perceived coping ability and psychiatric resilience in a dose-dependent manner (Table 3); every 1 SD unit increase in ACE-IQ scores was associated with lower levels of perceived coping ability (β=-0.14; 95% CI -0.15,-0.12) and psychiatric resilience (β=-0.28; 95% CI -0.29,-0.26) in the fully adjusted model. Associations between ACEs and perceived coping ability and psychiatric resilience were observed across levels of social support but were slightly stronger among women with low social support (Supplementary Table 4). Sensitivity analyses showed that the associations remained evident when excluding parental separation/divorce from the ACE-IQ score (Supplementary Table 5), and when excluding women with top/bottom 10% happiness values (Supplementary Table 6).

**Table 3.** Associations between the number of ACEs and perceived coping ability (CD-RISC) and psychiatric resilience (β and 95% CI)*

|  | N (%) | Perceived coping ability |  | Psychiatric resilience |  |
| --- | --- | --- | --- | --- | --- |
|  |  | Model 1<sup>a</sup> | Model 2<sup>b</sup> | Model 1<sup>a</sup> | Model 2<sup>b</sup> |
| **Number of ACEs** |  |  |  |  |  |
| 0 ACE | 4375 (22.31) | 0 (ref.) | 0 (ref.) | 0 (ref.) | 0 (ref.) |
| 1 ACE | 4485 (22.87) | -0.07 (-0.09, -0.05) | -0.06 (-0.07, -0.04) | -0.08 (-0.10, -0.06) | -0.07 (-0.09, -0.05) |
| 2 ACE | 3432 (17.50) | -0.10 (-0.11, -0.08) | -0.08 (-0.10, -0.07) | -0.13 (-0.14, -0.11) | -0.12 (-0.13, -0.10) |
| 3-4 ACE | 3979 (20.29) | -0.14 (-0.16, -0.13) | -0.12 (-0.14, -0.11) | -0.20 (-0.21, -0.18) | -0.18 (-0.20, -0.16) |
| ≥ 5 ACEs | 3342 (17.04) | -0.20 (-0.22, -0.18) | -0.16 (-0.18, -0.14) | -0.33 (-0.35, -0.31) | -0.29 (-0.31, -0.28) |

\*Coefficients are standardized; <sup>a</sup>adjusted for age and childhood deprivation; <sup>b</sup>additionally adjusted for education level, civil status, employment status and income

Poisson models revealed that compared to women with 0 ACEs, women with ≥ 5 ACEs had a lower prevalence of high perceived coping ability (PR=0.67, 95% CI 0.60,0.74), and high psychiatric resilience (PR=0.44, 95% CI 0.41,0.48) in the fully adjusted model (Table 4). Every unit increase in the ACE-IQ scores was associated with lower prevalence of high perceived coping ability (PR=0.94, 95% CI 0.92,0.95) and high psychiatric resilience (PR=0.87, 95% CI 0.86,0.88).

**Table 4.**
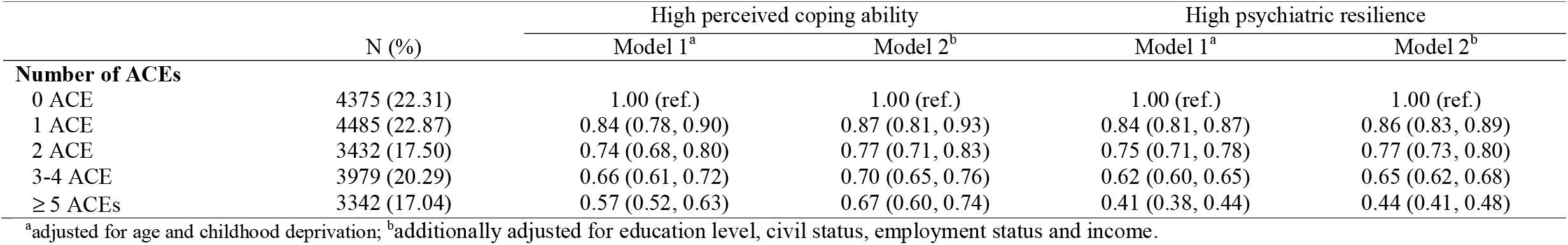
Prevalence Ratios (with 95% CI) of high perceived coping ability (CD-RISC ≥ 35) and high psychiatric resilience (absence of psychiatric morbidity) in relation to the number of ACEs

|  | N (%) | High perceived coping ability |  | High psychiatric resilience |  |
| --- | --- | --- | --- | --- | --- |
|  |  | Model 1<sup>a</sup> | Model 2<sup>b</sup> | Model 1<sup>a</sup> | Model 2<sup>b</sup> |
| **Number of ACEs** |  |  |  |  |  |
| 0 ACE | 4375 (22.31) | 1.00 (ref.) | 1.00 (ref.) | 1.00 (ref.) | 1.00 (ref.) |
| 1 ACE | 4485 (22.87) | 0.84 (0.78, 0.90) | 0.87 (0.81, 0.93) | 0.84 (0.81, 0.87) | 0.86 (0.83, 0.89) |
| 2 ACE | 3432 (17.50) | 0.74 (0.68, 0.80) | 0.77 (0.71, 0.83) | 0.75 (0.71, 0.78) | 0.77 (0.73, 0.80) |
| 3-4 ACE | 3979 (20.29) | 0.66 (0.61, 0.72) | 0.70 (0.65, 0.76) | 0.62 (0.60, 0.65) | 0.65 (0.62, 0.68) |
| ≥ 5 ACEs | 3342 (17.04) | 0.57 (0.52, 0.63) | 0.67 (0.60, 0.74) | 0.41 (0.38, 0.44) | 0.44 (0.41, 0.48) |

<sup>a</sup>adjusted for age and childhood deprivation; <sup>b</sup>additionally adjusted for education level, civil status, employment status and income.

ACE subtypes were associated with lower levels of perceived coping ability and psychiatric resilience (Supplementary Figure 2) as well as lower prevalence of high psychiatric resilience and high perceived coping ability except for physical neglect- and abuse, parental death or separation/divorce, and community- and collective violence (Supplementary Figure 3). After mutual adjustment for all ACE subtypes, we found that emotional neglect, being bullied, sexual abuse, and growing up with a mentally ill household member had consistent associations with both resilience measures, both in linear models (Figure 1) and Poisson models (Figure 2). Associations were also suggested for emotional abuse, domestic- and community violence with psychiatric resilience in the full model while associations for other ACEs were substantially attenuated (Figures 1 and 2).

**Figure 1.**
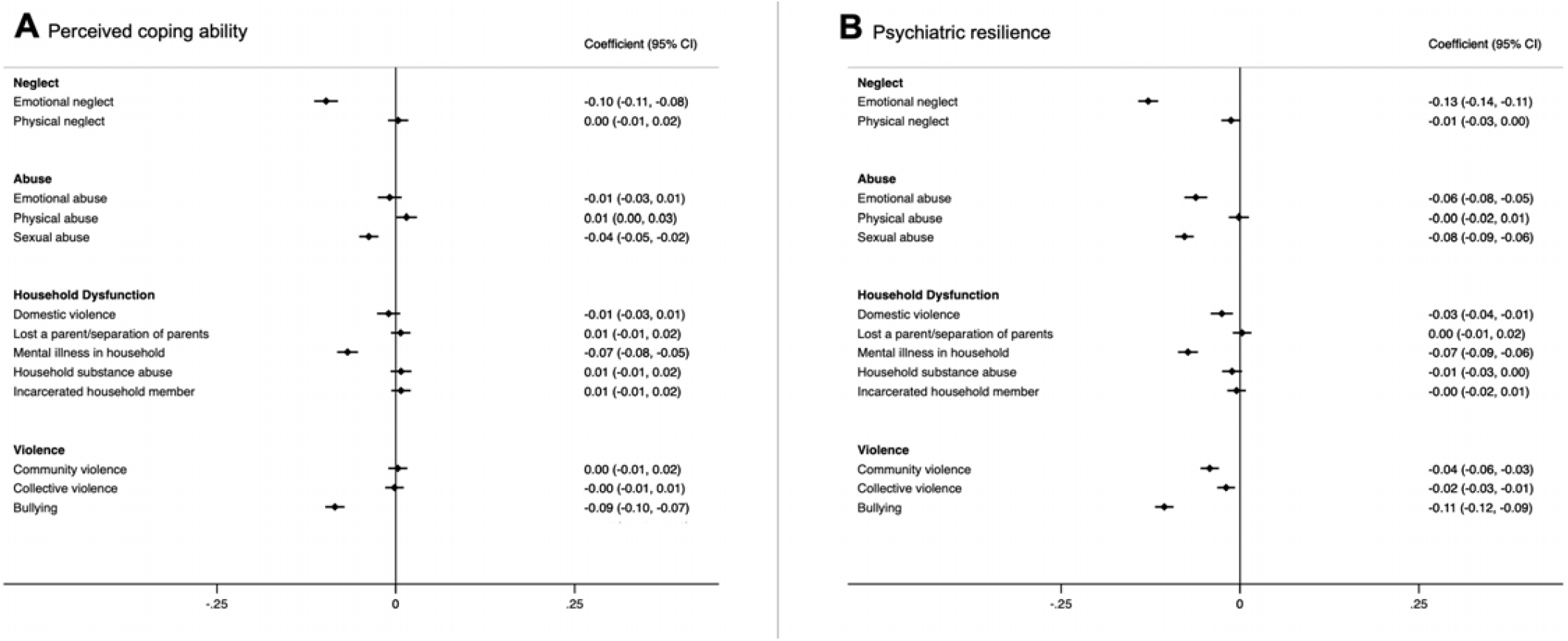
Associations between different types of ACEs and perceived coping ability (A) and psychiatric resilience (B) (β and 95% CI). Models were corrected for age, childhood deprivation, educational level, civil status, employment status, income and mutually adjusted for other ACEs. *Coefficients are standardized. Figure 2. Prevalence Ratios (with 95% CI) of high perceived coping ability (A) and high psychiatric resilience (B) in relation to individual ACEs. Models were corrected for age, childhood deprivation, educational level, civil status, employment status, income and mutually adjusted for other ACEs.

**Figure 2.**
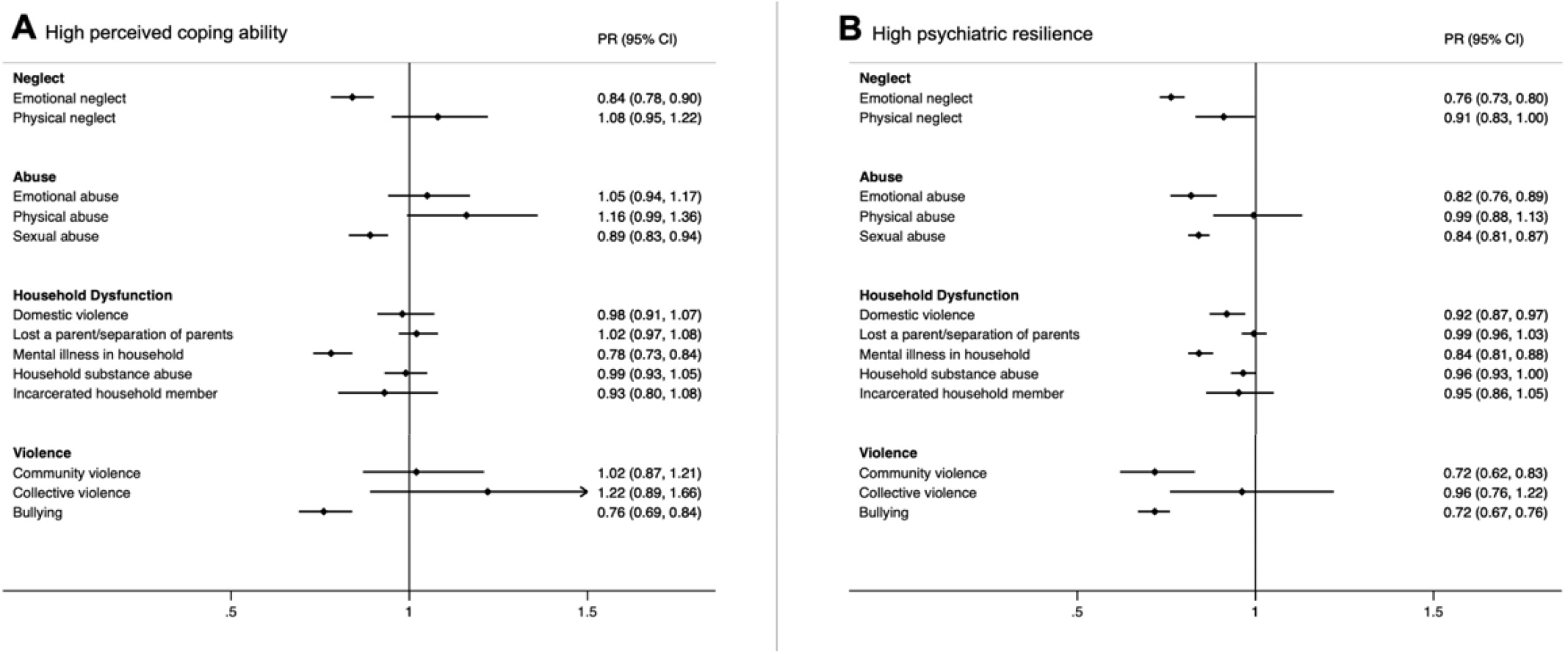
Prevalence Ratios (with 95% CI) of high perceived coping ability (A) and high psychiatric resilience (B) in relation to individual ACEs. Models were corrected for age, childhood deprivation, educational level, civil status, employment status, income and mutually adjusted for other ACEs.

## Discussion

In a large nationwide study of Icelandic women, a comprehensive measure of 13 adverse childhood experiences (ACEs) was negatively associated with two distinct measures of resilience in adulthood in a dose-dependent manner. Indeed, women who endorsed five or more ACEs had 33% lower prevalence of high perceived coping ability and 56% lower prevalence of high psychiatric resilience compared to women who endorsed 0 ACEs. In addition, specific ACEs including emotional neglect, bullying, sexual abuse and growing up with a mentally ill household member were strongly associated with attenuated adult resilience.

To our knowledge, the present study is the first to consider a wide range of ACEs in relation to adult resilience, and to address their association with two distinct resilience measures. Overall, our results are in line with previous studies examining the association between childhood maltreatment and adult resilience (Campbell-Sills et al., 2009; McGloin & Widom, 2001; Mersky & Topitzes, 2010; Nishimi, Choi, Davis, et al., 2020; Topitzes et al., 2013; Williams et al., 2006). However, the existing evidence base is limited by relatively small sample sizes, consideration of only few ACEs (Campbell-Sills et al., 2009; McGloin & Widom, 2001; Nishimi, Choi, Davis, et al., 2020; Williams et al., 2006), and/or the use of composite measures of childhood adversity (Mersky & Topitzes, 2010; Topitzes et al., 2013).

Our results indicating a dose-response relationship between the cumulative number of ACEs and attenuated adult resilience is consistent with a recent cross-sectional study by Nishimi et al. (2020) who observed a similar pattern between four ACEs (emotional abuse, physical abuse, sexual abuse, and witnessing domestic violence) and perceived coping ability, as measured with the CD-RISC. We have now extended these findings to psychiatric resilience as an outcome, and further demonstrated this pattern for a broad set of ACEs in a large nationwide study representing 30% of the Icelandic female population. Furthermore, our results suggest that all ACE subtypes are associated with perceived coping ability and psychiatric resilience. However, when mutually adjusting for other ACEs, only emotional neglect, sexual abuse, being bullied and growing up with a mentally ill household member were consistently associated with both resilience measures.

In line with previous literature (Nishimi, Choi, Cerutti, et al., 2020; Sheerin et al., 2018), we found that self-assessed coping ability and outcome-based psychiatric resilience were only moderately correlated with each other, indicating these may reflect different patterns of adaptive functioning following adversity. Collectively, our research adds to the growing literature on the nature of resilience as a construct. The associations between ACEs and adult resilience were independent of age, childhood deprivation and, importantly, other adult socio-demographic factors (i.e. educational level, civil status, employment level and income), suggesting that ACEs affect resilience over and above these potential mediating variables. In fact, effect sizes only diminished slightly when we additionally adjusted for adult socio-demographic factors, which indicates that adult characteristics such as education and employment level do not compensate for the deleterious impact of ACEs on adult resilience. This suggests that adult resilience may largely be determined in childhood and that situational factors in adulthood (e.g. high social support) only marginally buffer the association between ACEs and adult resilience.

The main strengths of the study include the population-based design and large sample size of the SAGA cohort, which represents the Icelandic adult female population in terms of distribution of age, residence, education and income (‘Preliminary findings of the Stress-And-Gene-Analysis Cohort’). The wealth of measures in the SAGA cohort baseline assessments made it possible to derive two types of resilience measures and examine a wide range of ACEs and take into account important covariates all of which have been limited in previous studies (Arseneault et al., 2011; Bellis et al., 2017; Copeland et al., 2018). However, our study also has several limitations. First, the cross-sectional nature of our data does not allow us to make any inferences about the directionality of the studied associations. Second, we cannot rule out that the observed association between ACE growing up with a mentally ill household member and resilience, is due to a genetic predisposition for psychopathology rather than the experience itself. Future genetically informative studies will need to examine the extent to which this association is confounded by genetic factors. In addition, ACEs were retrospectively reported and thus may be subject to recall bias. However, previous studies have shown acceptable validity for retrospective assessments of ACEs (Hardt & Rutter, 2004; Reuben et al., 2016; Widom & Morris, 1997; Widom & Shepard, 1996) although they may be influenced by current mental health status or response style (Reuben et al., 2016). Yet, the similar results obtained from our sensitivity analyses excluding individuals with extreme values on the happiness assessment, reduce concerns that our results are due to a particular response style. Finally, our results are based on an exclusively female sample, therefore, future studies should explore whether there are qualitative differences in how ACEs relate to adult resilience among men.

In conclusion, in a large nationwide-representative female population, we observed a strong association between cumulative exposure of ACEs and two distinct measures of adult resilience. Specific ACEs, including exposure to emotional neglect and bullying, sexual abuse, and growing up with a mentally ill household member were consistently associated with attenuated adult resilience. If these findings are confirmed through prospective designs, there may be huge societal benefits of prevention strategies targeting the protection of children against traumatic occurrences and their consequences. Future research is needed to address how children exposed to ACEs can be supported to reduce risks of compromised adult resilience and health inequalities.

## Data Availability

The data used in this study are compiled in the SAGA cohort and are available for review on site, on request. The use of these data is in accordance with the Icelandic law and with permission of the Icelandic Bioethics Committee. Therefore, the authors cannot make the dataset publicly available. On the other hand, interested researchers can obtain data access upon approvals by Icelandic Bioethics Committee (https://www.vsn.is/en) and by contacting the corresponding author.

## Ethics

The study was approved by the National Bioethics Committee (NBC number: 17-238) and all participants gave informed consent before participation.

