## Supplemental materials for "Adverse childhood experiences and resilience among adult women: a population-based study"

**Supplemental methods**

*Perceived coping ability*

The CD-RISC total scores were divided into quintiles which resulted in 18.61% of the sample in the lowest quintile (raw CD-RISC scores = 0 to 21), 20.00% in the lower middle quintile (raw CD-RISC scores = 22 to 26), 20.95% in the middle quintile (raw CD-RISC scores = 27 to 30), 19.13% in the higher middle quintile (raw CD-RISC scores = 31 to 34) and 21.31% in the highest quintile (raw CD-RISC scores = 35 to 40).

*Covariate assessment*

Age was divided into five groups for descriptive purposes: 18-29 years, 30-39 years, 40-49 years, 50-59 years, and 60 years and older. The age covariate was used as a continuous variable in models. Education was categorized as primary education, secondary education (high school or vocational education), tertiary education A (BSc or equivalent), and tertiary education B (MSc or above). Civil status was divided into married or in a relationship and single or widowed, and employment status was divided into employed (including being a student and being on parental leave) and retired or on disability or sick leave. Current monthly income was categorized into the following groups: low income (<$2527), low-medium income ($2528-$4212), medium income ($4213-$5897), medium-high income ($5898-$8424), and high income (>$8425; conversion rates according to Central Bank of Iceland, October 17, 2018). Childhood deprivation was assessed with the question: Was your family's economic situation ever so bad that you suffered any deprivation as a consequence? For example, this could apply to deprivation of nutritious food and/or deprivation of warm clothes and appropriate footwear during the winter months, with response options ranging from 0 (never) to 4 (often). Social support was assessed with the Multidimensional Scale of Perceived Social Support (MSPSS) (Zimet, Dahlem, Zimet, & Farley, 1988). The instrument consists of 12 items answered on a 7-point scale ranging from 0 (very strongly disagree) to 6 (very strongly agree). Items were summed to create a total score ranging from 0 to 72, with higher scores indicating higher levels of perceived social support. In addition, the total scores were divided into quartiles which resulted in 25.09% of the sample in the lowest quartile (raw MSPSS scores = 0 to 49), 25.52% of the sample in the low-middle quartile (raw MSPSS scores = 50 to 61), 24.92% of the sample in the high-middle quartile (raw MSPSS scores = 62 to 69) and 24.47% of the sample in the highest quartile (raw MSPSS scores = 70 to 72). A categorical variable was then created where the highest tertile was used to define a high level of social support, the two middle quartiles were merged and used together to define a moderate level of social support and the lowest quartile was used to define a low level of social support. Happiness was assessed with the question "In general, how would you rate your happiness?", and participants rated their happiness with a slider ranging from 1 to 10.

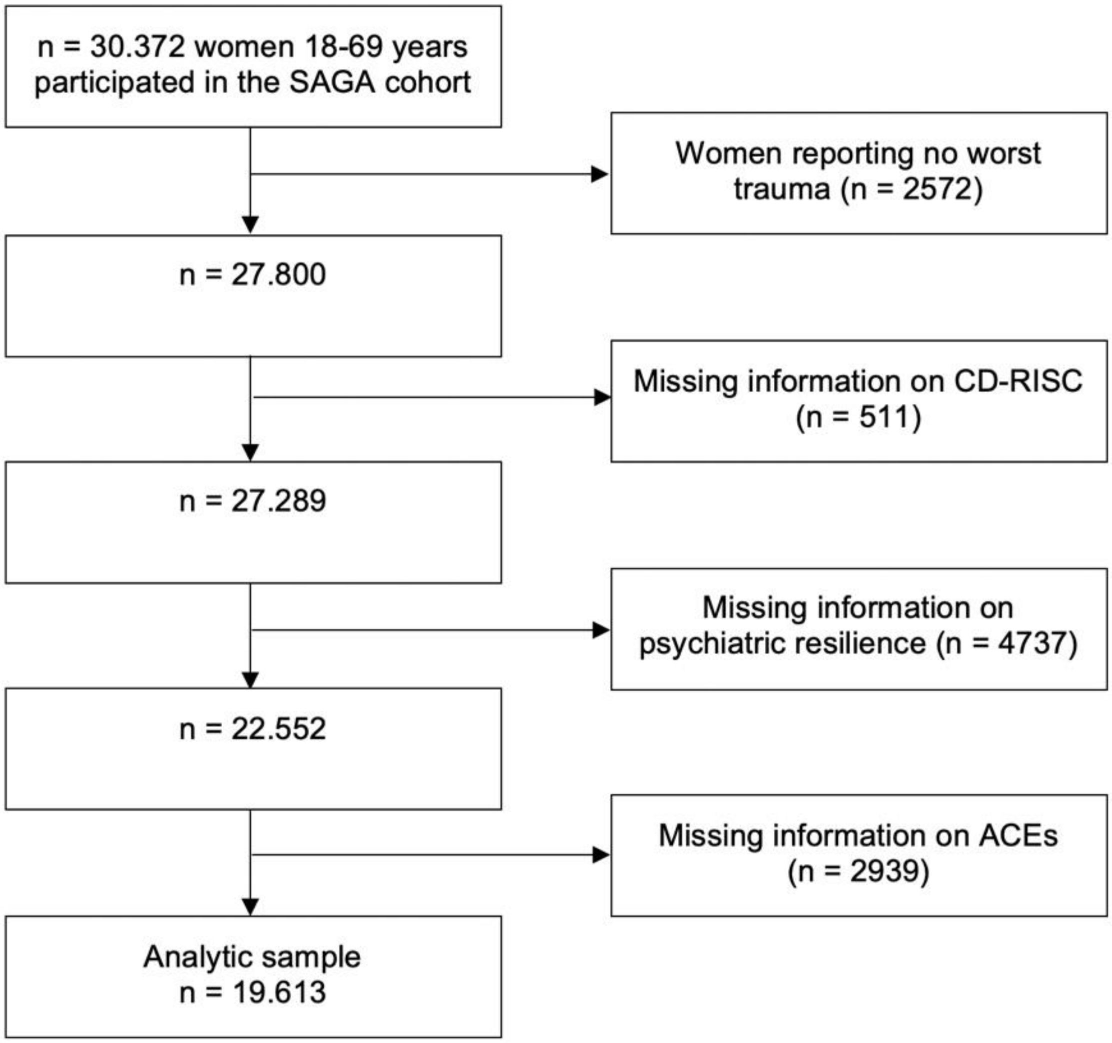


Supplementary Figure 1. Flow-chart of the study population

Supplementary Table 1. List of the 30 ACE-IQ items used to derive the 13 different ACEs and their response options.

| ACE item | Scoring* |
|---|---|
| ***Neglect*** | |
| **Emotional neglect** | |
| Did your parents/guardians understand your problems and worries? | Always=0, Most of the time=1, Sometimes=2, Rarely=3, Never=4 |
| Did your parents/guardians really know what you were doing with your free time when you were not at school or work? | Always=0, Most of the time=1, Sometimes=2, Rarely=3, Never=4 |
| **Physical neglect** | |
| How often did your parents/guardians not give you enough food even when they could easily have done so? | Never=0, Once=1, A few times=2, Many times=3 |
| Were your parents/guardians too drunk or intoxicated by drugs to take care of you? | Never=0, Once=1, A few times=2, Many times=3 |
| How often did your parents/guardians not send you to school even when it was available? | Never=0, Once=1, A few times=2, Many times=3 |
| ***Abuse*** | |
| **Emotional abuse** | |
| Did a parent, guardian or other household member yell, scream or swear at you, insult or humiliate you? | Never=0, Once=1, A few times=2, Many times=3 |
| Did a parent, guardian or other household member threaten to, or actually, abandon you or throw you out of the house? | Never=0, Once=1, A few times=2, Many times=3 |
| **Physical abuse** | |
| Did a parent, guardian or other household member spank, slap, kick, punch or beat you up? | Never=0, Once=1, A few times=2, Many times=3 |
| Did a parent, guardian or other household member hit or cut you with an object, such as a stick (or cane), bottle, club, knife, whip etc? | Never=0, Once=1, A few times=2, Many times=3 |
| **Sexual abuse** | |
| Did someone touch or fondle you in a sexual way when you did not want them to? | Never=0, Once=1, A few times=2, Many times=3 |
| Did someone make you touch their body in a sexual way when you did not want them to? | Never=0, Once=1, A few times=2, Many times=3 |
| Did someone attempt oral, anal, or vaginal intercourse with you when you did not want them to? | Never=0, Once=1, A few times=2, Many times=3 |
| Did someone actually have oral, anal, or vaginal intercourse with you when you did not want them to? | Never=0, Once=1, A few times=2, Many times=3 |
| ***Household dysfunction*** | |
| **Domestic violence** | |
| Did you see or hear a parent or household member in your home being yelled at, screamed at, sworn at, insulted or humiliated? | Never=0, Once=1, A few times=2, Many times=3 |
| Did you see or hear a parent or household member in your home being slapped, kicked, punched or beaten up? | Never=0, Once=1, A few times=2, Many times=3 |
| Did you see or hear a parent or household member in your home being hit or cut with an object, such as a stick (or cane), bottle, club, knife, whip etc.? | Never=0, Once=1, A few times=2, Many times=3 |

Supplementary Table 1 (continued)

| | |
|---|---|
| **Lost a parent / separation of parents** | |
| Were your parents ever separated or divorced? | No=0, Yes=1 |
| Did your mother, father or guardian die? | No=0, Yes=1 |
| **Mental illness in household** | |
| Did you live with a household member who was depressed, mentally ill or suicidal? | No=0, Yes=1 |
| **Household substance abuse** | |
| Did you live with a household member who was a problem drinker or alcoholic, or misused street or prescription drugs? | No=0, Yes=1 |
| **Incarcerated household member** | |
| Did you live with a household member who was ever sent to jail or prison? | No=0, Yes=1 |
| ***Other violence*** | |
| **Community violence** | |
| Did you see or hear someone being beaten up in real life? | Never=0, Once=1, A few times=2, Many times=3 |
| Did you see or hear someone being stabbed or shot in real life? | Never=0, Once=1, A few times=2, Many times=3 |
| Did you see or hear someone being threatened with a knife or gun in real life? | Never=0, Once=1, A few times=2, Many times=3 |
| **Collective violence** | |
| During the first 18 years of your life, were you exposed to war/collective violence (e.g. from gangs or police)?** | No=0, Yes=1 |
| Were you forced to go and live in another place due to any of these events? | Never=0, Once=1, A few times=2, Many times=3 |
| Did you experience the deliberate destruction of your home due to any of these events? | Never=0, Once=1, A few times=2, Many times=3 |
| Were you beaten up by soldiers, police, militia, or gangs? | Never=0, Once=1, A few times=2, Many times=3 |
| Was a family member or friend killed or beaten up by soldiers, police, militia, or gangs? | Never=0, Once=1, A few times=2, Many times=3 |
| **Bullying** | |
| How often were you bullied? | Never=0, Once=1, A few times=2, Many times=3 |

*all items also had the option „can‘t/don‘t want to answer“

**this is a screening question, only participants that responded yes got the following four questions

### Supplemental results

Supplementary Table 2. Rank order correlations for perceived coping (CD-RISC) and different measures of psychopathology used to derive the psychiatric resilience phenotype (n =19,613)[a]

| | CD-RISC | PHQ-9 | GAD-7 | PCL-5 | PSQI-A | Binge drinking |
|---|---|---|---|---|---|---|
| CD-RISC | 1 | | | | | |
| PHQ-9 | -0.53 | 1 | | | | |
| GAD-7 | -0.50 | 0.76 | 1 | | | |
| PCL-5 | -0.47 | 0.68 | 0.65 | 1 | | |
| PSQI-A | -0.40 | 0.61 | 0.62 | 0.62 | 1 | |
| Binge drinking | -0.05 | 0.12 | 0.12 | 0.06 | 0.08 | 1 |

Supplementary Table 3. Rank order correlations for ACE subtypes (n =19,613)[a]

| | Emotional abuse | Physical abuse | Sexual abuse | Emotional neglect | Physical neglect | Domestic violence | Lost a parent/ separation | Mental illness in household | Household substance abuse | Incarcerated household member | Community violence | Collective violence | Bullying |
|---|---|---|---|---|---|---|---|---|---|---|---|---|---|
| Emotional abuse | 1 | | | | | | | | | | | | |
| Physical abuse | 0.46 | 1 | | | | | | | | | | | |
| Sexual abuse | 0.17 | 0.13 | 1 | | | | | | | | | | |
| Emotional neglect | 0.35 | 0.22 | 0.21 | 1 | | | | | | | | | |
| Physical neglect | 0.28 | 0.17 | 0.13 | 0.26 | 1 | | | | | | | | |
| Domestic violence | 0.51 | 0.31 | 0.17 | 0.33 | 0.28 | 1 | | | | | | | |
| Lost a parent/separation of parents | 0.15 | 0.09 | 0.12 | 0.18 | 0.18 | 0.20 | 1 | | | | | | |
| Mental illness in household | 0.31 | 0.17 | 0.14 | 0.25 | 0.22 | 0.36 | 0.18 | 1 | | | | | |
| Household substance abuse | 0.20 | 0.11 | 0.14 | 0.23 | 0.27 | 0.35 | 0.25 | 0.30 | 1 | | | | |
| Incarcerated household member | 0.14 | 0.10 | 0.08 | 0.11 | 0.17 | 0.21 | 0.14 | 0.17 | 0.25 | 1 | | | |
| Community violence | 0.20 | 0.20 | 0.09 | 0.13 | 0.14 | 0.17 | 0.07 | 0.11 | 0.10 | 0.11 | 1 | | |
| Collective violence | 0.10 | 0.09 | 0.06 | 0.06 | 0.07 | 0.08 | 0.05 | 0.07 | 0.06 | 0.09 | 0.10 | 1 | |
| Bullying | 0.20 | 0.15 | 0.12 | 0.15 | 0.12 | 0.14 | 0.06 | 0.15 | 0.08 | 0.06 | 0.12 | 0.08 | 1 |

Supplementary Table 4. Associations between the number of ACEs and perceived coping ability (CD-RISC) and psychiatric resilience stratified by social support (n=19,398) (β and 95% CI)

| | | Perceived coping ability | | | Psychiatric resilience | | |
|---|---|---|---|---|---|---|---|
| | N (%) | Low support | Moderate support | High support | Low support | Moderate support | High support |
| **Number of ACEs** | | | | | | | |
| 0 ACE | 4340 (22.37) | 0 (ref.) | 0 (ref.) | 0 (ref.) | 0 (ref.) | 0 (ref.) | 0 (ref.) |
| 1 ACE | 4445 (22.91) | -0.07 (-0.11, -0.02) | -0.05 (-0.07, -0.03) | -0.03 (-0.05, -0.00) | -0.10 (-0.14, -0.06) | -0.06 (-0.09, -0.04) | -0.05 (-0.07, -0.03) |
| 2 ACE | 3399 (17.52) | -0.09 (-0.14, -0.05) | -0.06 (-0.08, -0.04) | -0.06 (-0.09, -0.04) | -0.13 (-0.18, -0.09) | -0.10 (-0.13, -0.08) | -0.09 (-0.12, -0.07) |
| 3-4 ACE | 3939 (20.31) | -0.12 (-0.16, -0.08) | -0.09 (-0.11, -0.07) | -0.07 (-0.09, -0.04) | -0.21 (-0.25, -0.17) | -0.16 (-0.18, -0.14) | -0.11 (-0.14, -0.09) |
| ≥ 5 ACEs | 3275 (16.88) | -0.17 (-0.21, -0.13) | -0.09 (-0.12, -0.07) | -0.07 (-0.10, -0.03) | -0.32 (-0.36, -0.28) | -0.25 (-0.28, -0.23) | -0.20 (-0.24, -0.17) |

*Coefficients are standardized; [a]adjusted for age and childhood deprivation; [b]additionally adjusted for education level, civil status, employment status and income

Supplementary Table 5. Associations between the number of ACEs (excluding parental divorce/separation) and perceived coping ability (CD-RISC) and psychiatric resilience (β and 95% CI)*

| | | Perceived coping ability | | Psychiatric resilience | |
|---|---|---|---|---|---|
| | N (%) | Model 1[a] | Model 2[b] | Model 1[a] | Model 2[b] |
| **Number of ACEs** | | | | | |
| 0 ACE | 5158 (26.30) | 0 (ref.) | 0 (ref.) | 0 (ref.) | 0 (ref.) |
| 1 ACE | 4941 (25.19) | -0.08 (-0.10, -0.06) | -0.07 (-0.08, -0.05) | -0.09 (-0.11, -0.07) | -0.08 (-0.09, -0.06) |
| 2 ACE | 3646 (18.59) | -0.11 (-0.13, -0.10) | -0.10 (-0.11, -0.08) | -0.15 (-0.17, -0.14) | -0.15 (-0.16, -0.13) |
| 3-4 ACE | 3814 (19.45) | -0.17 (-0.18, -0.15) | -0.14 (-0.16, -0.13) | -0.24 (-0.25, -0.22) | -0.22 (-0.23, -0.20) |
| ≥ 5 ACEs | 2054 (10.47) | -0.18 (-0.20, -0.16) | -0.14 (-0.15, -0.12) | -0.30 (-0.32, -0.29) | -0.27 (-0.29, -0.25) |

*Coefficients are standardized; [a]adjusted for age and childhood deprivation; [b]additionally adjusted for education level, civil status, employment status and income

Supplementary Table 6. Associations between the number of ACEs and perceived coping ability (CD-RISC) and psychiatric resilience excluding participants with ≈10% lowest and highest happiness values (raw scores 1-5 and 10) (n=15,449) (β and 95% CI)*

| | | Perceived coping ability | | Psychiatric resilience | |
|---|---|---|---|---|---|
| | N (%) | Model 1[a] | Model 2[b] | Model 1[a] | Model 2[b] |
| **Number of ACEs** | | | | | |
| 0 ACE | 3499 (22.65) | 0 (ref.) | 0 (ref.) | 0 (ref.) | 0 (ref.) |
| 1 ACE | 3594 (23.26) | -0.06 (-0.08, -0.04) | -0.04 (-0.06, -0.03) | -0.08 (-0.10, -0.06) | -0.07 (-0.09, -0.05) |
| 2 ACE | 2803 (18.14) | -0.08 (-0.10, -0.06) | -0.06 (-0.08, -0.05) | -0.12 (-0.14, -0.10) | -0.11 (-0.13, -0.09) |
| 3-4 ACE | 3155 (20.42) | -0.11 (-0.13, -0.09) | -0.09 (-0.11, -0.07) | -0.17 (-0.19, -0.15) | -0.16 (-0.18, -0.14) |
| ≥ 5 ACEs | 2398 (15.52) | -0.14 (-0.16, -0.12) | -0.11 (-0.13, -0.09) | -0.29 (-0.31, -0.27) | -0.27 (-0.28, -0.25) |

*Coefficients are standardized; [a]adjusted for age and childhood deprivation; [b]additionally adjusted for education level, civil status, employment status and income

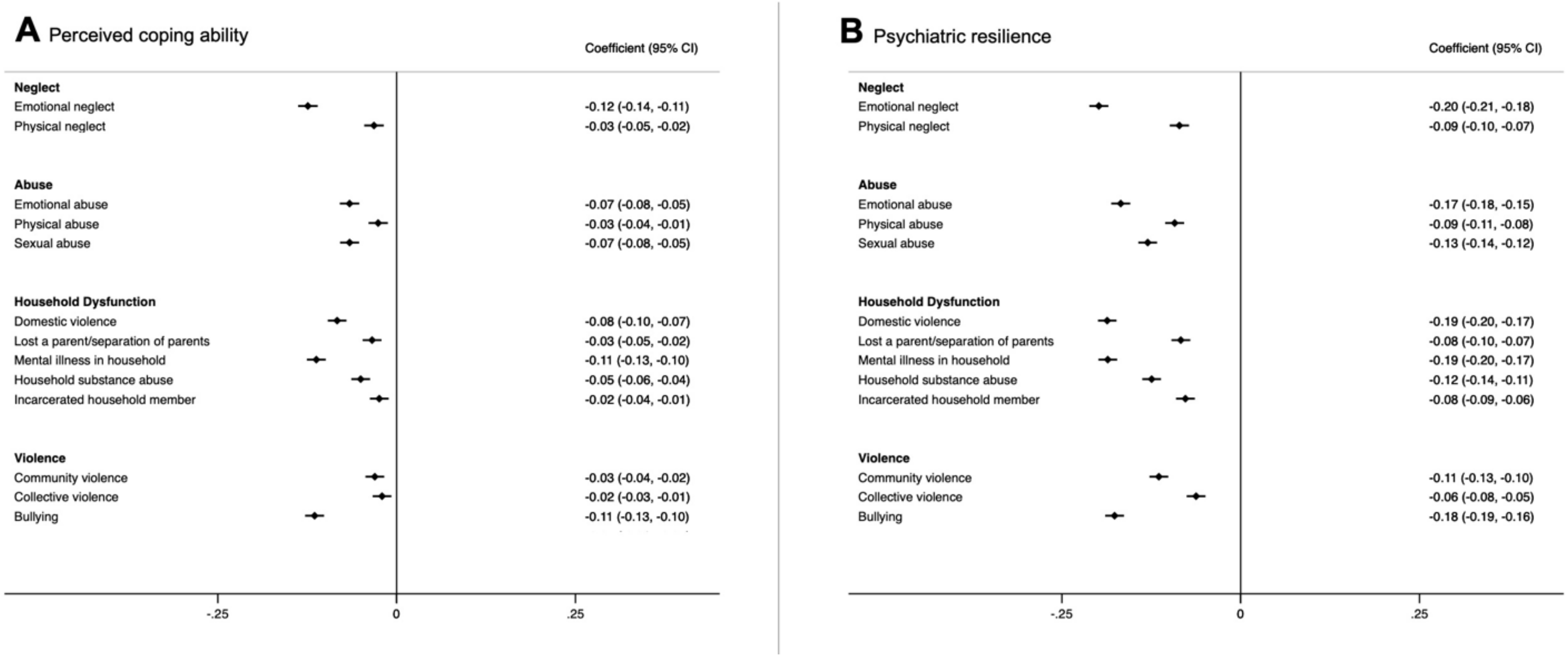


Supplementary Figure 1. Associations between different types of ACEs and perceived coping ability (A) and psychiatric resilience (B) (β and 95% CI). Models were corrected for age, childhood deprivation, education level, civil status, employment status and income.
*Coefficients are standardized.

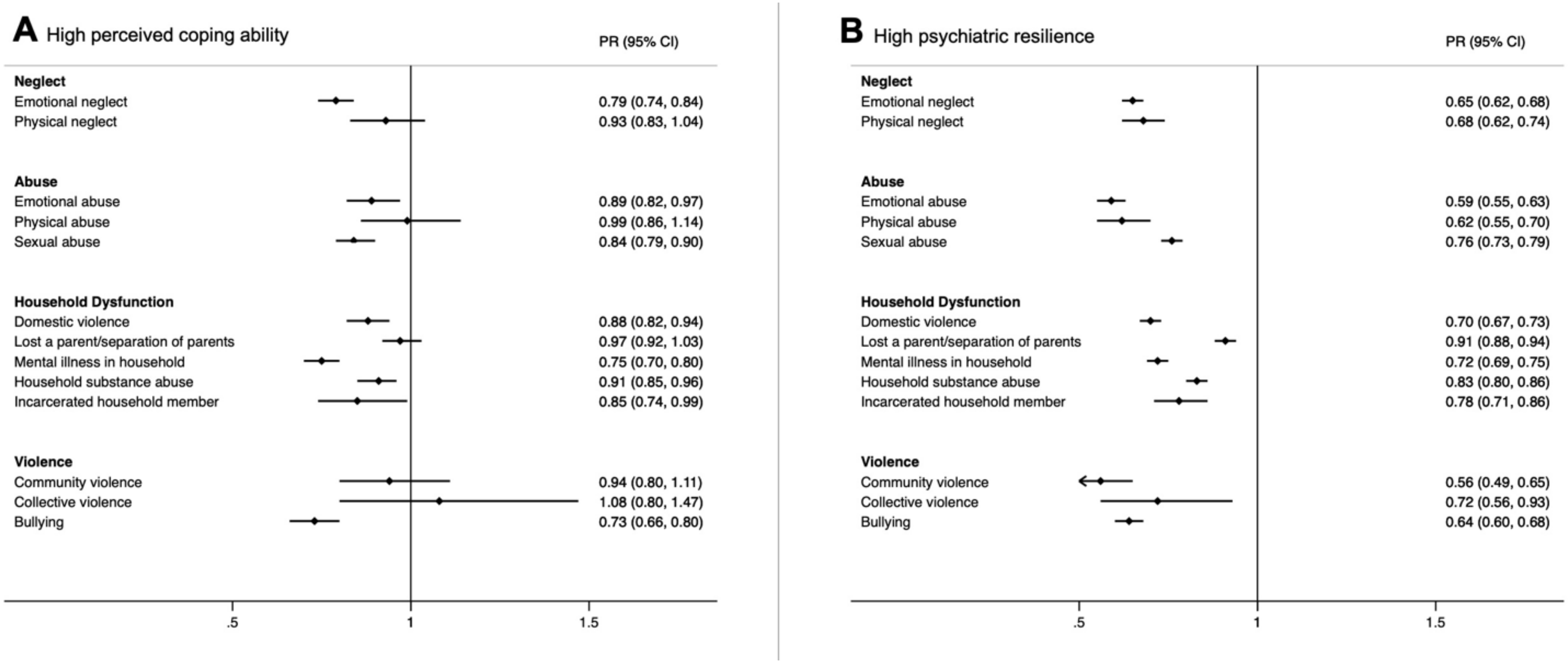


Supplementary Figure 3. Prevalence Ratios (with 95% CI) of high perceived coping ability (A) and high psychiatric resilience (B) in relation to individual ACEs. Models were corrected for age, childhood deprivation, education level, civil status, employment status and income.
